# Effective Heat Inactivation of SARS-CoV-2

**DOI:** 10.1101/2020.04.29.20085498

**Authors:** Tony T. Wang, Christopher Z. Lien, Shufeng Liu, Prabhuanand Selvaraj

**Affiliations:** Laboratory of Vector-Borne Viral Diseases, Division of Viral Products, Center for Biologics Evaluation, U.S. Food and Drug Administration, Silver Spring, Maryland, United States of America

## Abstract

In this study, we aimed to evaluate the stability of SARS-CoV-2 under four different heat conditions (37, 42, 56, 60 °C) and report that the virus is stable at 37 °C for at least 24 hours. Heating at 56 °C for 30 minutes, however, effectively inactivated the virus while preserved the stability of viral RNA in both human sera and sputum samples. These findings provide critical information regarding the biology of the virus as well as a practical way to inactivate infectious virus that is potentially found in clinical specimens.

## Manuscript text

The outbreak of the severe acute respiratory syndrome coronavirus 2 (SARS-CoV-2 or 2019-nCoV) has quickly turned into a global pandemic^1^. Infectious viruses had been isolated from oro- or naso-pharyngeal swabs^2^, sputum^2^ and possibly stool samples^3^ of infected individuals. Handling these clinical specimens therefore poses a biosafety risk to both healthcare professionals and laboratory workers. This study aimed to determine the effect of various heat treatment conditions on the infectivity and RNA stability of SARS-CoV-2 with the goal to identify a practical approach to inactivate the virus in clinical specimens.

SARS-CoV-2 stocks with an infectious titer of 1.4×10^7^ TCID50/ml were heated at four different temperatures for 15, 30, 60, 90, 120 minutes. The infectivity was then determined by a TCID50 assay based on the Reed & Muench method. Shown in Table 1, heating at 37 °C for up to 2 hours failed to effectively reduce the infectivity. Virus appeared to be stable at 37 °C for at least 24 hours with only marginal decrease in infectivity and then became much less infectious after 48 hours heat treatment (reduction by 6 logs). By contrast, heating at 60°C for just 15 minutes completely inactivated the virus (reduced infectivity by more than 7 logs). At 42 °C, heating up to 60 minutes only minimally reduced viral infectivity. At 56 °C, virus infectivity decreased by about 3–4 logs after 15 minutes and went below detection limit after 30 minutes (reduction by 7 logs). Furthermore, the presence of 50% human serum does not alter the inactivation rate.

**Table 1.** Inactivation of SARS-CoV-2 under different heat conditions.

| Temperature | Virus titers (TCID50/ml) |  |  |  |  |  |  |
| --- | --- | --- | --- | --- | --- | --- | --- |
|  | 15 min | 30 min | 60 min | 90 min | 120 min | 24hrs | 48hrs |
| 37°C | 5.6x10^6 | 1.0x10^7 | 2.5x10^7 | 1.8x10^7 | 2.5x10^7 | 1.8x10^6 | 320 |
| 42°C | 3.2x10^5 | 3.2x10^5 | 1.9X10^6 | n.d.* |  | 1-10 | undetectable |
| 56°C | 2.5x10^3 | undetectable | undetectable |  |  | n.d.* |  |
| 56°C (50% human serum) | n.d.* | undetectable | n.d.* |  |  |  |  |
| 60°C | undetectable | undetectable | undetectable |  |  |  |  |
| Unheated | 1.4x10^7 |  |  |  |  |  |  |

\*n.d., not determined

To mimic clinical specimens, we spiked various amounts of virus into sputum samples collected from five healthy donors. We then split each sputum in two halves. One set of samples were heat inactivated at 56°C for 30 minutes, and the other set was left untreated. Following standard viral isolation using the Qiagen QIAamp viral RNA isolation kit, viral RNA was quantified by real-time PCR. Again, there was no loss of RNA in sputum samples that were heated at 56°C for 30 minutes (Table 2). Given 56 °C is more commonly used in serology laboratories, we further quantified the amount of viral RNA in spiked sera samples (containing 50% human serum) prior to and after heat-inactivation. In consistent, the virus RNA levels remain comparable before and after heating at 56 °C for 30 minutes.

**Table 2.** Heat-treatment at 56°C for 30 minutes does not reduce viral RNA in specimens^*^.

|  | Viral loads (GCE/μl) (Ct value) |  |  |  |  |  |  |
| --- | --- | --- | --- | --- | --- | --- | --- |
| 56°C<br>30 min | Sputum 1 | Sputum 2 | Sputum 3 | Sputum 4 | Sputum 5 | 50% human serum 1 | 50% human serum 2 |
| Before | 22882 (23.4) | 1705 (26.7) | 64 (31) | 6.9 (33.9) | 2.0 (35.9) | 37534.9 | 159.8 |
| After | 18924 (23.6) | 2038 (26.5) | 65 (31) | 6.6 (33.9) | 2.5 (35.1) | 44524.5 | 191.2 |

GCE, Genome Copy Equivalent; Ct, cycle threshold
\*Viral RNA was extracted from 140 μl spiked specimens (sputum or serum). 5 out of 60 μl of total RNA was used in real-time PCR. Results were plotted as GCE/μl of RNA.

Although coronaviruses are generally thermolabile, SARS-CoV-2 appears to be stable at 37°C for at least 24 hours. This finding may help to explain why some countries with a tropical climate have not been able to escape from SARS-CoV-2. Heating specimens at 56 °C for 30 minutes, whether in the presence of human serum or in sputum, effectively inactivated any infectious virus in the samples while preserved the viral RNA. This conclusion is robust because we tested undiluted virus stock with infectious virus titer to the 10^7^ TCID50 as well as dilutions nearly to the detection limit of the real-time PCR assay. It is unlikely any clinical specimens containing detectible amount of infectious virus would go beyond the range that we have studied. Therefore, we conclude that heating clinical samples at 56 °C for 30 minutes may be a practical way for healthcare and laboratory professionals to eliminate biosafety risks associated with processing clinical specimens of COVID-19 patients.

Limitations of this study include: no humidity factor was evaluated and the lack of large numbers of clinical specimens for statistical analysis.

## Data Availability

Data available.

## Acknowledgements

The following reagent was deposited by the Centers for Diseases Control and Prevention and obtained through BEI Resources, NIAID, NIH: SARS-Related Coronavirus 2, Isolate USA-WA1/2020, NR-52281.

## Notes

### Competing Interest Statement

The authors have declared no competing interest.

### Funding Statement

Funded by the U.S. Food and Drug Administration.

## References

1. WHO. WHO Director-General’s opening remarks at the media briefing on COVID-19. 2020.

2. Wolfel R, Corman VM, Guggemos W, et al. Virological assessment of hospitalized patients with COVID-2019. Nature. 2020.

3. Zhang Y CC, Zhu S et al. Isolation of 2019-nCoV from a stool specimen of a laboratory-confirmed case of the coronavirus disease 2019 (COVID-19). China CDC Weekly. 2020;2(8):123–124.

